# Machine learning models for the prediction of COVID-19 prognosis in the primary health care setting

**DOI:** 10.1101/2025.05.08.25327245

**Authors:** Joan Barrot, Joan A. Caylà, Manel Mata-Cases, Jordi Real, Bogdan Vlacho, Josep Franch-Nadal, Didac Mauricio, the COVID-19 Working Group in Primary Health Care

## Abstract

**Objective:** This study aimed to identify prognostic factors associated with poor outcomes of COVID-19 at diagnosis in Primary Health Care (PHC).

**Methods:** We conducted a retrospective, longitudinal study using the SIDIAP database, part of the PHC Information System of Catalonia. The analysis included COVID-19 cases diagnosed in patients aged 18 and older from March 2020 to September 2022. Follow-up was conducted for 90 days post-diagnosis or until death. Various machine learning models of differing complexities were used to predict short-term events, including mortality and hospital complications. Each model was tailored to maximize the predictive accuracy for poor outcomes, exploring algorithms such as Generalized Linear Models, flexible GLMs with Lasso, Gradient Boosting Models, and Support Vector Machines, with the model demonstrating the highest Area Under the Curve (AUC) selected for optimal performance.

**Results:** A total of 2,162,187 COVID-19 cases were identified across five epidemic waves. Key predictors of short-term complications included age and the epidemic wave. Additional significant factors encompassed social deprivation (MEDEA), blood pressure, cardiovascular history, chronic obstructive pulmonary disease (COPD), obesity, and diabetes mellitus. The models exhibited high performance, with AUC values ranging from 0.73 to 0.95. A web application was developed to estimate the risk of adverse outcomes based on individual patient profiles (https://dapcat.shinyapps.io/CovidScore).

**Conclusions:** In addition to age and epidemic wave, predictors such as social deprivation, diabetes mellitus, obesity, COPD, cardiovascular disease, high blood pressure, and dyslipidemia significantly indicate poor prognosis in COVID-19 patients diagnosed in PHC, and the developed application facilitates risk quantification for individual patients.

## Introduction

The COVID-19 pandemic has posed unprecedented challenges to healthcare systems globally. Since its identification in December 2019 in Wuhan, China, this virus has spread rapidly, affecting millions and causing an international global health crisis [1]. Until December 2022, more than 704 million cases and more than 6.7 million deaths were reported [2]. In this context, the best strategy to control this pandemic has been discussed (Zero COVID-19 vs. mitigation) [3]. However, in most healthcare settings, primary healthcare (PHC) has played a crucial role in the response to the pandemic [4]. In Europe, the first cases of COVID-19 were detected in January 2020, reaching a peak in this first wave on March 31; Spain had a cumulative incidence of 178.03/1 million inhabitants, followed by Italy with 92.66 and Austria with 61.14 [2]. The first cases in Spain were detected in February 2020, [5], and a state of alarm was declared on March 15, 2020, which lasted until June 21, 2020. During this period, the Ministry of Health was in charge of the protocols and organization of healthcare related to COVID-19 nationwide [6].

PHC was instrumental in detecting, monitoring, and treating patients and their contacts worldwide. The role of PHC varied significantly between countries, and the different health system models influenced it. There were two main challenges for PHC: 1) to clearly define its role in providing an effective contribution to the prevention, diagnosis, and treatment of COVID-19 and, thus, reorganize accordingly; 2) to identify new ways of providing regular health services and maintaining the quality of care for non-COVID-19 patients [7].

The evolution of COVID-19 has shown considerable variability in the risk of severe complications and mortality, depending on the individual characteristics of the patients [8]. Known risk factors include high blood pressure, diabetes mellitus, chronic lung diseases, and obesity. Other prognostic factors may vary depending on the epidemic’s different phases and the population’s vaccination status [8,9]. Some factors are present during diagnosis, while others can be identified through complementary analysis or tests.

Guidance to support general practitioners in managing future waves of COVID-19 or other health emergencies should be tailored to general practice from the outset. Establishing risk factors associated with severity and prognosis in the early stages of the disease is important to identify patients who need specialized care as a priority. For this reason, creating new clinical tools to improve health decisions and outcomes in the population is essential.

Our study aimed to develop machine learning models to identify prognostic factors and predict the outcome in subjects with COVID-19 during diagnosis in PHC settings.

## Material and Methods

### General Design

We conducted a retrospective cohort study using data from the PHC Information System of Catalonia (SIDIAP) database, which includes the healthcare data from individuals from all PHC centers and hospitals within the Catalan Health System (CatSalut), the main healthcare provider in Catalonia, Northeast Spain. The study was conducted according to the guidelines of the Declaration of Helsinki, and approved by the Institutional Review Board (or Ethics Committee) of IDIAP Jordi Gol i Gurina Foundation (code 21/271-PCV). Data were accessed for research purposes on June 30th 2021. Authors did not had access to information that could identify individual participants during or after data collection.

### Study selection criteria

During the observational period (the initial two and half years of the pandemic, 03/2020-09/2022), we included all individuals in the database with a positive COVID-19 diagnostic test or diagnostic code (ICD-10: B34.2; B97.2; B97.21; B97.29; J12.81; J12.89; U07.1; Z20.828; J12.89; J20.8; J22; J40; J80; J98.8). We only selected subjects aged 18 years or older with at least one year of data as a registered user in the SIDIAP database. During the study, each episode of COVID-19 infection in the same patient was considered as an independent COVID-19 case.

### Study variables

Variables available in PHC at the time of identification and/or diagnosis of each COVID-19 patient were evaluated as potential predictors. These included sociodemographic factors: age, sex, the MEDEA deprivation index (Mortality in small Spanish areas and Socioeconomic and Environmental Inequalities) [10], toxic habits (e.g., smoking), vaccination status, and the infection period (pandemic wave). Clinically relevant medical conditions were identified by the presence of ICD-10 diagnostic codes for diabetes mellitus, cardiovascular disease, obesity, ischemic heart disease, stroke, heart failure, peripheral arterial disease, chronic kidney disease (CKD), hypertension, dyslipidemia, diabetic retinopathy, diabetic neuropathy, dementia, asthma, COPD, obstructive sleep apnea syndrome and/or use of CPAP, deep vein thrombosis, HIV and malignancies. We collected the following clinical variables closest to the inclusion date, within a window of one year prior to inclusion: HbA1c, body mass index (BMI), blood pressure, lipid profile, aspartate transaminase (AST) and alanine transaminase (also known as alanine aminotransferase) (ALT), estimated glomerular filtration rate (eGFR), urine albumin to creatinine ratio (ACR), C reactive protein (CRP), leukocytes, leukocyte formula, hemoglobin, ferritin, platelets, lactate dehydrogenase (LDH), and ferritin levels.

### Study events

The follow-up period was defined as the time from the date of COVID-19 diagnosis to a maximum of 90 days after inclusion or until death. We assessed the following events: death from any cause, hospitalization, admission to the intensive care unit (ICU), and complications attributed to COVID-19, including the need for mechanical ventilation, as well as respiratory, neurological, thrombotic, and cardiovascular complications.

### Statistical analysis

A descriptive analysis was performed on the 2,162,187 identified COVID-19 cases across five epidemic waves. To optimize the modeling process and avoid unnecessary use of computational resources, a random sample of 100,000 cases was selected from the initial dataset for training and testing. The learning curves for the selected models were generated [11], showing that the AUC for the test and training samples stabilized and converged after approximately 40,000 cases (see learning curves in Supplementary Figure 1).

The model construction process included: data preprocessing, selection of training (75%) and validation (25%) samples, selection of the machine learning (ML) algorithm with the best performance, and the type of ML of different levels of complexity: The types of algorithms tested included generalized polynomial models (GLM), flexible GLM with Lasso (elastic net regularization), gradient boosting models (GBM), and support vector machine (SVM) models. The model chosen was the one that demonstrated the best performance, based on the AUC criteria and following the parsimony principle by selecting only the most important features for each model. The process also involved adjusting, validating, optimizing, and selecting the most optimal model, followed by its implementation within an integrated application. The initial variables included for the models followed semi-agnostic criteria, prioritizing those with known clinical associations with the outcomes under study. All models were designed to include sociodemographic variables such as age, sex, and epidemic wave. For continuous variables with potential missing values (excluding age), quantile-based categorization strategies that included a ‘missing’ category were used to account for incomplete data.

Finally, the selected model was implemented in a Shiny-based APP (https://dapcat.shinyapps.io/CovidScore), where the risk of each event was calculated for each associated model, with graphical representations of risk levels and the importance of each variable. The entire process of data management, statistical analysis, and model construction was carried out using the R Core Team (2022) software [12], along with the shiny R package [13] for the implementation of the models. The XAI (Explainable AI) methodology was applied to improve the transparency and interpretability of the machine learning models using the DALEX (v2.4.3) R package [14]. This package offers graphical tools that provide model-agnostic explainers for exploring and understanding complex predictive models, such as neural networks and ensembles. These explainers facilitate the decomposition of predictions, performance assessment, and model comparison, thereby aiding in the validation and interpretation of predictive ‘black boxes’. The main R packages utilized for the modeling were stats [15], GLMnet [16], gbm [17], E1071 [18], and caret [11].

## Results

### Evolution of COVID-19 cases and epidemic waves

A total of 2,162,187 cases of COVID-19 were identified between March 2020 and September 2022 in 5 epidemic waves. A total of 242,692 (11.2%) cases were diagnosed in the first wave, 736,020 (34.0%) in the second, 329,230 (15.2%) in the third, 199,606 (9.23%) in the fourth, and 654,639 (30.3%) in the fifth. (**Figure 1**).

**Figure 1.**
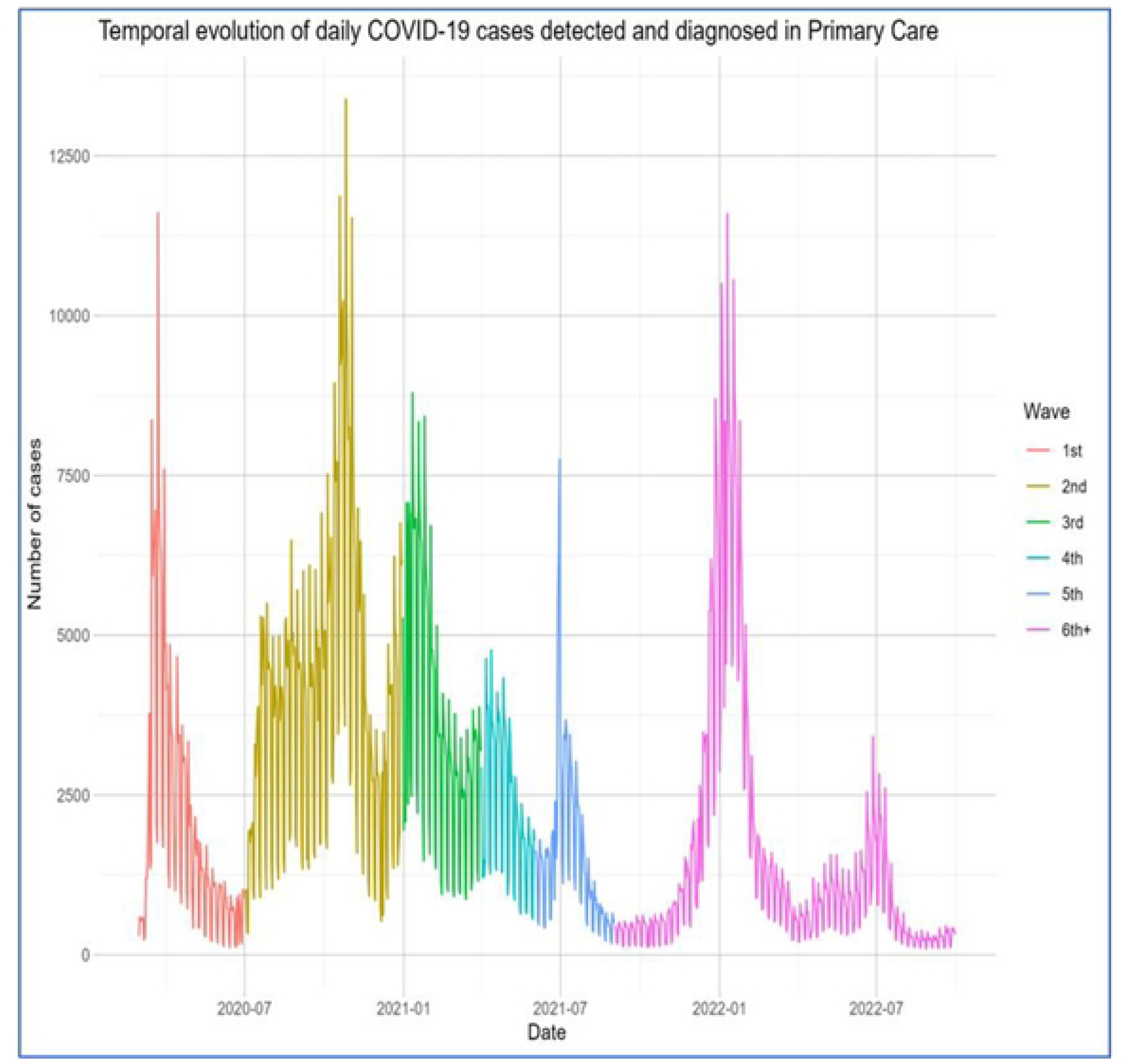
Graphical representation of the evolution of COVID-19 cases in five epidemic waves in Catalonia from March 2020 to September 2022.

### Characteristics of COVID-19 cases

The study population had a mean age of 46.5 years (standard deviation [SD] = 18.5), and 56.5% were female. Among the participants, 24.6% were smokers, and 32.8% had received at least one dose of a COVID-19 vaccine. The proportion of vaccinated individuals increased across successive waves. Additional sociodemographic characteristics and clinical variables, overall and by wave, are detailed in **Table 1**.

**Table 1:**
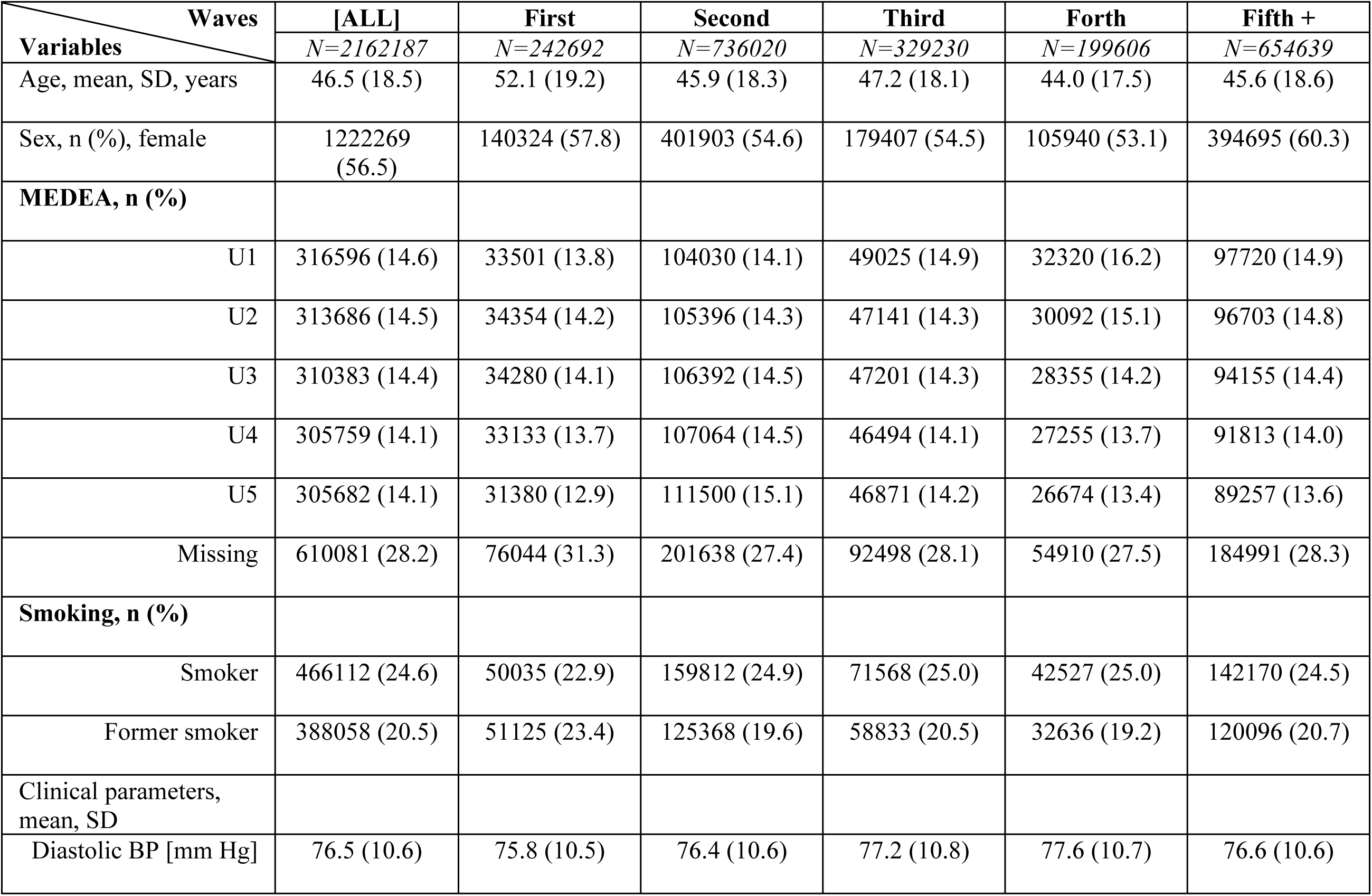
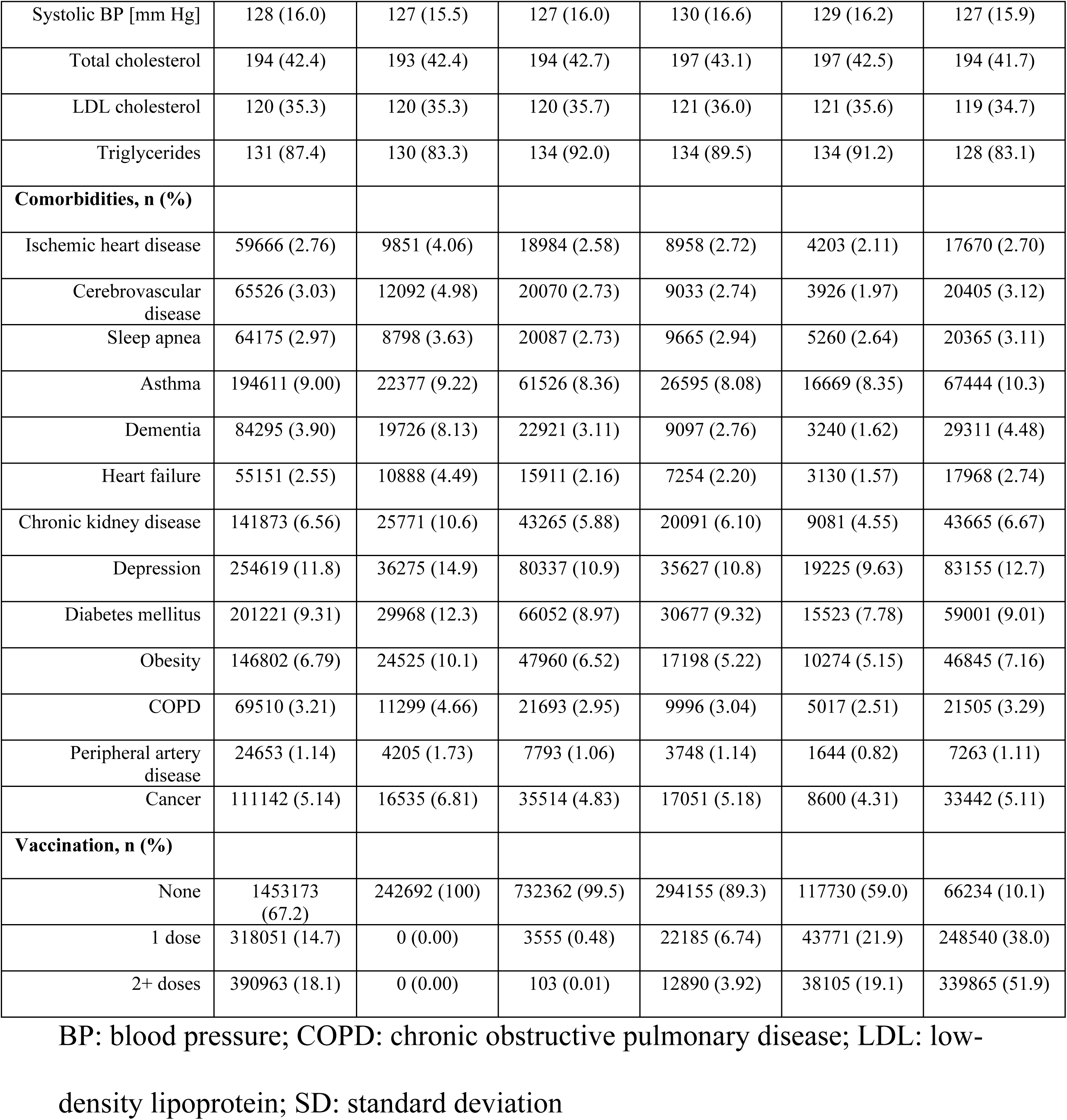
Characteristics of the population in each epidemic wave.

| Waves | [ALL] | First | Second | Third | Forth | Fifth + |
| --- | --- | --- | --- | --- | --- | --- |
| Variables | N=2162187 | N=242692 | N=736020 | N=329230 | N=199606 | N=654639 |
| Age, mean, SD, years | 46.5 (18.5) | 52.1 (19.2) | 45.9 (18.3) | 47.2 (18.1) | 44.0 (17.5) | 45.6 (18.6) |
| Sex, n (%), female | 1222269 (56.5) | 140324 (57.8) | 401903 (54.6) | 179407 (54.5) | 105940 (53.1) | 394695 (60.3) |
| MEDEA, n (%) |  |  |  |  |  |  |
| U1 | 316596 (14.6) | 33501 (13.8) | 104030 (14.1) | 49025 (14.9) | 32320 (16.2) | 97720 (14.9) |
| U2 | 313686 (14.5) | 34354 (14.2) | 105396 (14.3) | 47141 (14.3) | 30092 (15.1) | 96703 (14.8) |
| U3 | 310383 (14.4) | 34280 (14.1) | 106392 (14.5) | 47201 (14.3) | 28355 (14.2) | 94155 (14.4) |
| U4 | 305759 (14.1) | 33133 (13.7) | 107064 (14.5) | 46494 (14.1) | 27255 (13.7) | 91813 (14.0) |
| U5 | 305682 (14.1) | 31380 (12.9) | 111500 (15.1) | 46871 (14.2) | 26674 (13.4) | 89257 (13.6) |
| Missing | 610081 (28.2) | 76044 (31.3) | 201638 (27.4) | 92498 (28.1) | 54910 (27.5) | 184991 (28.3) |
| Smoking, n (%) |  |  |  |  |  |  |
| Smoker | 466112 (24.6) | 50035 (22.9) | 159812 (24.9) | 71568 (25.0) | 42527 (25.0) | 142170 (24.5) |
| Former smoker | 388058 (20.5) | 51125 (23.4) | 125368 (19.6) | 58833 (20.5) | 32636 (19.2) | 120096 (20.7) |
| Clinical parameters, mean, SD |  |  |  |  |  |  |
| Diastolic BP [mm Hg] | 76.5 (10.6) | 75.8 (10.5) | 76.4 (10.6) | 77.2 (10.8) | 77.6 (10.7) | 76.6 (10.6) |
| Systolic BP [mm Hg] | 128 (16.0) | 127 (15.5) | 127 (16.0) | 130 (16.6) | 129 (16.2) | 127 (15.9) |
| Total cholesterol | 194 (42.4) | 193 (42.4) | 194 (42.7) | 197 (43.1) | 197 (42.5) | 194 (41.7) |
| LDL cholesterol | 120 (35.3) | 120 (35.3) | 120 (35.7) | 121 (36.0) | 121 (35.6) | 119 (34.7) |
| Triglycerides | 131 (87.4) | 130 (83.3) | 134 (92.0) | 134 (89.5) | 134 (91.2) | 128 (83.1) |
| **Comorbidities, n (%)** |  |  |  |  |  |  |
| Ischemic heart disease | 59666 (2.76) | 9851 (4.06) | 18984 (2.58) | 8958 (2.72) | 4203 (2.11) | 17670 (2.70) |
| Cerebrovascular disease | 65526 (3.03) | 12092 (4.98) | 20070 (2.73) | 9033 (2.74) | 3926 (1.97) | 20405 (3.12) |
| Sleep apnea | 64175 (2.97) | 8798 (3.63) | 20087 (2.73) | 9665 (2.94) | 5260 (2.64) | 20365 (3.11) |
| Asthma | 194611 (9.00) | 22377 (9.22) | 61526 (8.36) | 26595 (8.08) | 16669 (8.35) | 67444 (10.3) |
| Dementia | 84295 (3.90) | 19726 (8.13) | 22921 (3.11) | 9097 (2.76) | 3240 (1.62) | 29311 (4.48) |
| Heart failure | 55151 (2.55) | 10888 (4.49) | 15911 (2.16) | 7254 (2.20) | 3130 (1.57) | 17968 (2.74) |
| Chronic kidney disease | 141873 (6.56) | 25771 (10.6) | 43265 (5.88) | 20091 (6.10) | 9081 (4.55) | 43665 (6.67) |
| Depression | 254619 (11.8) | 36275 (14.9) | 80337 (10.9) | 35627 (10.8) | 19225 (9.63) | 83155 (12.7) |
| Diabetes mellitus | 201221 (9.31) | 29968 (12.3) | 66052 (8.97) | 30677 (9.32) | 15523 (7.78) | 59001 (9.01) |
| Obesity | 146802 (6.79) | 24525 (10.1) | 47960 (6.52) | 17198 (5.22) | 10274 (5.15) | 46845 (7.16) |
| COPD | 69510 (3.21) | 11299 (4.66) | 21693 (2.95) | 9996 (3.04) | 5017 (2.51) | 21505 (3.29) |
| Peripheral artery disease | 24653 (1.14) | 4205 (1.73) | 7793 (1.06) | 3748 (1.14) | 1644 (0.82) | 7263 (1.11) |
| Cancer | 111142 (5.14) | 16535 (6.81) | 35514 (4.83) | 17051 (5.18) | 8600 (4.31) | 33442 (5.11) |
| **Vaccination, n (%)** |  |  |  |  |  |  |
| None | 1453173 (67.2) | 242692 (100) | 732362 (99.5) | 294155 (89.3) | 117730 (59.0) | 66234 (10.1) |
| 1 dose | 318051 (14.7) | 0 (0.00) | 3555 (0.48) | 22185 (6.74) | 43771 (21.9) | 248540 (38.0) |
| 2+ doses | 390963 (18.1) | 0 (0.00) | 103 (0.01) | 12890 (3.92) | 38105 (19.1) | 339865 (51.9) |

BP: blood pressure; COPD: chronic obstructive pulmonary disease; LDL: low-
density lipoprotein; SD: standard deviation

### Incidence of complications post COVID-19

The descriptive analysis of incidence rates per event, particularly in the context of waves, shows a progressive reduction in case fatality rate (CFR) (overall and hospital), hospital admission, and different complications over time (**Table 2**). The highest incidence rate was for respiratory complications, at 7.4% (excluding composite endpoint non-fatal complications), while the lowest was for thrombotic complications, affecting only 0.15% of the population.

**Table 2:**
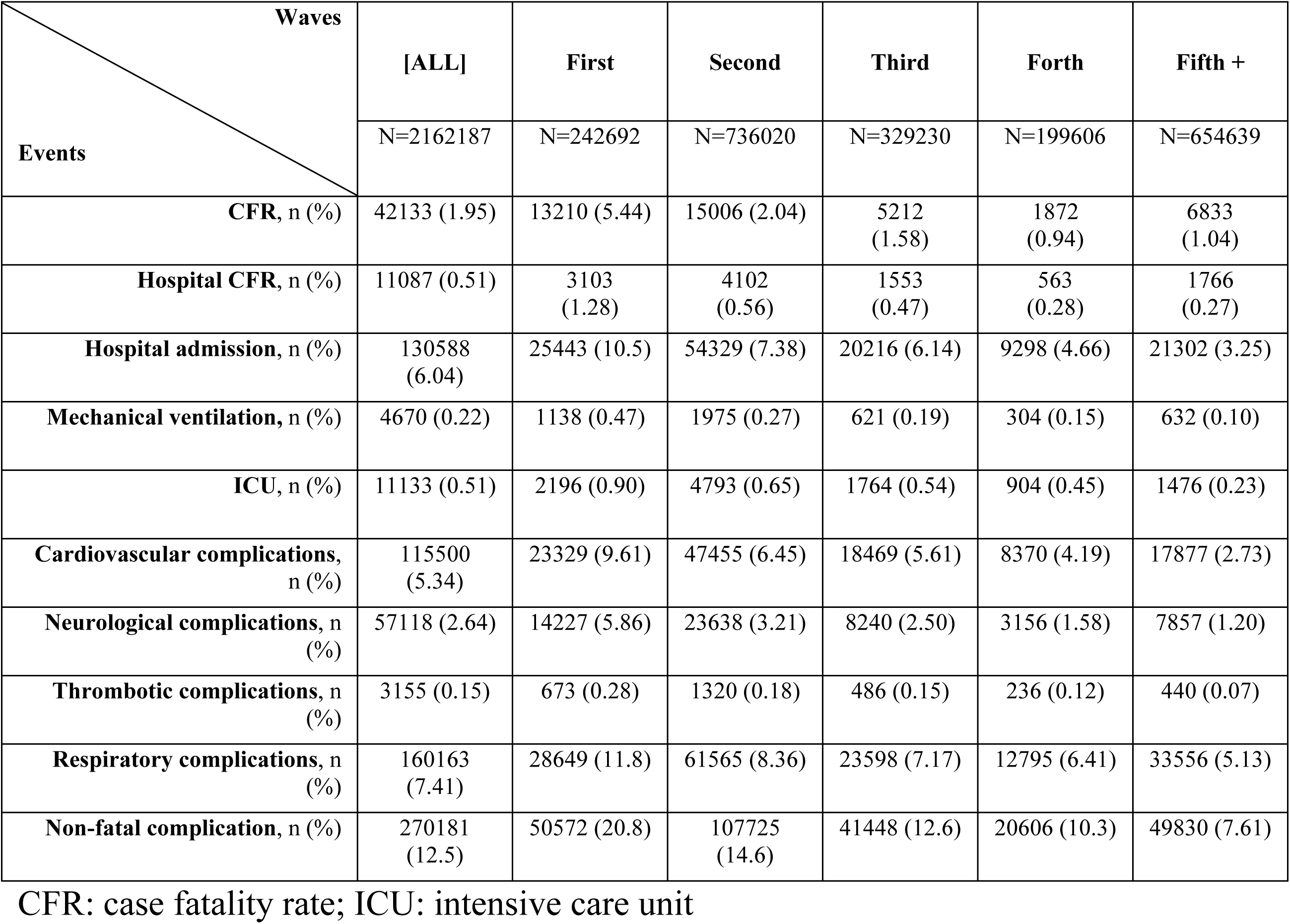
Complications for each epidemic wave from March 2020 to September 2022.

| Events \ Waves | [ALL] | First | Second | Third | Forth | Fifth + |
| --- | --- | --- | --- | --- | --- | --- |
|  | N=2162187 | N=242692 | N=736020 | N=329230 | N=199606 | N=654639 |
| CFR, n (%) | 42133 (1.95) | 13210 (5.44) | 15006 (2.04) | 5212 (1.58) | 1872 (0.94) | 6833 (1.04) |
| Hospital CFR, n (%) | 11087 (0.51) | 3103 (1.28) | 4102 (0.56) | 1553 (0.47) | 563 (0.28) | 1766 (0.27) |
| Hospital admission, n (%) | 130588 (6.04) | 25443 (10.5) | 54329 (7.38) | 20216 (6.14) | 9298 (4.66) | 21302 (3.25) |
| Mechanical ventilation, n (%) | 4670 (0.22) | 1138 (0.47) | 1975 (0.27) | 621 (0.19) | 304 (0.15) | 632 (0.10) |
| ICU, n (%) | 11133 (0.51) | 2196 (0.90) | 4793 (0.65) | 1764 (0.54) | 904 (0.45) | 1476 (0.23) |
| Cardiovascular complications, n (%) | 115500 (5.34) | 23329 (9.61) | 47455 (6.45) | 18469 (5.61) | 8370 (4.19) | 17877 (2.73) |
| Neurological complications, n (%) | 57118 (2.64) | 14227 (5.86) | 23638 (3.21) | 8240 (2.50) | 3156 (1.58) | 7857 (1.20) |
| Thrombotic complications, n (%) | 3155 (0.15) | 673 (0.28) | 1320 (0.18) | 486 (0.15) | 236 (0.12) | 440 (0.07) |
| Respiratory complications, n (%) | 160163 (7.41) | 28649 (11.8) | 61565 (8.36) | 23598 (7.17) | 12795 (6.41) | 33556 (5.13) |
| Non-fatal complication, n (%) | 270181 (12.5) | 50572 (20.8) | 107725 (14.6) | 41448 (12.6) | 20606 (10.3) | 49830 (7.61) |

CFR: case fatality rate; ICU: intensive care unit

### Predictors according to the selected model

The predictor variables selected for different events and the type of model are summarized in **Supplementary Table 1**. The models showed high performance, with the AUC ranging from 0.73 to 0.95. We found that the predictors for short-term complications with greater weight common to all models were age and epidemic wave. Other common predictors in our analysis were the deprivation index (MEDEA), blood pressure, presence of cardiovascular disease, COPD, diabetes, obesity, or chronic kidney disease. The mortality model had the highest performance with an AUC of 0.95, while the respiratory model had the lowest at 0.73. This and other performance metrics for each event model can be seen in **Supplementary Table 2**.

### Implementation of models

The models integrated into the APP-Web model can be accessed through this link: (https://dapcat.shinyapps.io/CovidScore). Based on a patient’s profile, this app estimates the risk for each endpoint (e.g., mortality, hospital admission, cardiovascular complications, etc., as shown in **Supplementary Figure 2**). **Supplementary Figure 2** presents a bar plot of the app’s main screen, illustrating an example patient profile and the corresponding output, which includes a bar plot showing the risk estimations for each endpoint. Additionally, the app visually displays the breakdown profile, overall model performance, feature importance, and the local performance of each model for individual patient profiles.

### Breakdown profile

For each risk estimate (e.g., mortality, hospital admission, cardiovascular complications, etc.), the app provides details of the contribution of each factor to the final probability calculation (see Breakdown profile in https://dapcat.shinyapps.io/CovidScore, with an example for the CV outcome shown in **Supplementary Figure 3**). The breakdown profile plot decomposes the estimated risk for each prediction. For example, for the risk estimate of cardiovascular complications, the plot provides detailed insights into the contribution of each factor to the final probability calculation. For a 97-year-old male with chronic kidney disease, being 97 years old increases the risk of 9.5%, being male increases the risk of 1.7%, and the presence of diabetes increases the risk of 4.2%, respectively, to the final estimated risk of 34%.

### Feature-importance and ROC curve by model

In **Supplementary Figure 4**, predictors for the mortality model are displayed and ranked according to their relative importance (feature importance) as determined by the model, along with the ROC curve comparing the training sample to the test sample. In the mortality model, the most important features were the COVID wave, sex, and neoplasia condition. The ROC curve showed good discrimination in both the test and training samples. The results of feature importance and ROC curves for the other endpoints can be seen on the app web.

## Discussion

This retrospective study analyzed over 2 million COVID-19 cases from March 2020 to September 2022 across multiple COVID-19 waves, with follow-up 90 days post-diagnosis or until death. Our study underscores the significant impact of COVID-19 on PHC in the initial years of this pandemic in Catalonia. Furthermore, using machine learning models (GLMs, Lasso, Gradient Boosting, SVMs), we identified key predictors of poor outcomes, such as age, social deprivation (MEDEA), blood pressure, and a history of either diabetes, COPD, cardiovascular disease, or obesity. The models showed strong predictive accuracy (AUC: 0.73–0.95). Finally, using these models, an interactive web app was developed for personalized risk estimation (https://dapcat.shinyapps.io/CovidScore).

We found that the CFR was highest during the first wave of the pandemic, gradually decreasing in subsequent waves, with the second wave showing the highest incidence of cases. The decrease in the CFR may be attributed to a combination of increased immunity (due to vaccination and SARS-CoV-2 infections), better identification of more severe cases, and the lower pathogenicity of recent variants like Omicron. Overall, these findings suggest changes in the SARS-CoV-2 virus, an adaptive response in healthcare, and improvements in the prevention (i.e., via vaccination) and treatment of complications as the pandemic progressed [19].

Regarding predictors of poor prognosis at the time of COVID-19 diagnosis in PHC, we identified older age, epidemic wave, social deprivation, and a history of diabetes, obesity, chronic obstructive pulmonary disease (COPD), cardiovascular disease, hypertension, and dyslipidemia. While study results vary across different populations, numerous studies have identified advanced age and comorbidities such as hypertension, cardiovascular disease, COPD, and diabetes as predictors of increased COVID-19 severity [20–22]. Additionally, social deprivation indicators, such as the MEDEA index, have been associated with poorer outcomes in terms of severity and mortality, underscoring the multifactorial nature of COVID-19 outcomes. Understanding and addressing these predictive factors is crucial for improving management and outcomes in affected patients.

Conditions associated with low-grade chronic inflammation, such as obesity and diabetes mellitus, are also relevant at the metabolic level [23]. Several systematic reviews have provided consistent evidence that diabetes and obesity are associated with poorer COVID-19 outcomes, which agrees with our study [24]. Although the reasons for this association are not entirely clear, these conditions could exacerbate respiratory problems and/or affect immune responses. A systematic review and meta-analyses on high-risk phenotypes in people with diabetes determined that individuals with a more severe course of diabetes and pre-existing comorbidities had a poorer prognosis of COVID-19 than individuals with a milder course of the disease, highlighting the need for individualized and proactive management strategies for high-risk patients [25]. At the hospital level, predictive models like the ISARIC 4C have been developed to anticipate clinical deterioration (including mortality, ICU admission, or intubation), assessing age, gender, comorbidities, and nosocomial infection [26]. A similar study in the United Kingdom, utilizing computerized PHC medical records, developed predictive algorithms for COVID-19 mortality and hospital admission risk. Factors such as age, body weight, ethnicity, and social risk explained 73% of COVID-19 deaths and 58% of hospital admissions, suggesting periodic recalibration of these models to reflect the evolving nature of the pandemic [27]. An early pandemic study on PHC identified key risk factors for ICU admission and mortality, including advanced age, male gender, autoimmune disease, bilateral pulmonary infiltrates, and elevated LDH, D-dimer, and C-reactive protein. Protective factors included myalgias, arthralgias, and anosmia [28]. Recent advances in AI and machine learning have significantly contributed to managing the COVID-19 pandemic by aiding in detection, treatment, mortality prediction, and infection modeling to reduce virus spread [29–31].

In this study, we used machine learning to develop predictive models and then used these models to develop an app. The app provides comprehensive information to estimate the risk of COVID-19 prognosis outcomes for individuals based on their risk factors (e.g., age, sex, comorbidities, vaccination status, COVID-19 wave). This approach could be used in PHC to identify individuals needing closer monitoring and interventions to prevent serious complications and hospitalization.

Our study has limitations inherent to its retrospective design. Outcomes depend on the quality of existing clinical records not specifically collected for this research and have yet to undergo individual validation. A notable limitation is the potential impact of vaccine implementation on epidemiology and prognosis, which underscores the need to recalibrate predictive models with post-vaccination data to maintain accuracy. Periodic updates with the latest available data are essential to ensure the continued relevance of these models. Although no predictive model is perfect for COVID-19 patients, our models serve as valuable tools to estimate the risk of complications, helping to identify patients who require closer monitoring. However, the accuracy of these models can be affected by variations in the detection and recording of symptoms and risk factors by different healthcare professionals. Moreover, mild COVID-19 cases may have gone unrecorded in PHC, potentially leading to their exclusion from our study population. However, the study benefits from using the SIDIAP database, which includes a substantial patient cohort and is a well-validated source for epidemiological and pharmaco-epidemiological studies within the Catalan primary care setting. This database not only provides standardized clinical data (including health issues, physical exams, lab results, and medication records) from pseudo-anonymized electronic health records but was also specifically updated to include COVID-19-related variables (such as diagnostic tests and procedures), enabling researchers to conduct targeted epidemiological studies.

## Conclusions

This study highlights the importance of different prognostic factors, including age, epidemic wave, and comorbidities, in assessing the risk of mortality and complications in patients with COVID-19 treated in PHC. Identifying these predictors is crucial to optimizing medical care and highlights the need for further research and recalibration of predictive models as epidemiological circumstances evolve, such as vaccination. Integrating these models into clinical practice will enable healthcare professionals to make informed and personalized decisions, thereby improving outcomes for patients affected by COVID-19. The developed application allows a fast risk quantification for each patient seen in PHC centers.

## Statements

### Use of artificial intelligence tools

None declared.

### Data availability

The data analyzed in this study is subject to the following licenses/restrictions: restrictions apply to the availability of some, or all data generated or analyzed during this study because they were used under license. The corresponding author will, on request, detail the restrictions and any conditions under which access to some data may be provided. Requests to access these datasets should be directed to Dr Bogdan Vlacho PharmD, MSc, PhD, at.

### Preprint

None

### Authors’ contributions

**J.B; J.A.C, M.M-C, D. M, J. F-N and J.R** conceptualized and designed the study**; J.R** conducted statistical analysis and data management; **B. V** contributed to data acquisition**; J.B; J.A.C, D. M, J. F-N, B.V and J.R** edited and cross-reviewed the manuscript. All authors approved the final version of the manuscript. **J.B** and **J.A.C,** contributed equally to this work and share first authorship.

